# The association of age-related and off-target retention with longitudinal quantification of [^18^F]MK6240 tau-PET in target regions

**DOI:** 10.1101/2022.05.24.22275386

**Authors:** Cécile Tissot, Stijn Servaes, Firoza Lussier, João Pedro Ferrari Souza, Joseph Therriault, Pâmela Cristina Lukasewicz Ferreira, Gleb Bezgin, Bruna Bellaver, Douglas Teixeira Leffa, Sulantha S. Mathotaarachchi, Jenna Stevenson, Nesrine Rahmouni, Min Su Kang, Vanessa Pallen, Nina Margherita-Poltronetti, Yi-Ting Wang, Jaime Fernandez-Arias, Andrea L. Benedet, Eduardo R. Zimmer, Jean-Paul Soucy, Dana L. Tudorascu, Annie D. Cohen, Madeleine Sharp, Serge Gauthier, Gassan Massarweh, Brian Lopresti, William E. Klunk, Suzanne L. Baker, Victor L. Villemagne, Pedro Rosa-Neto, Tharick A. Pascoal

**Author notes:** Corresponding author: Tharick A Pascoal, MD PhD, Assistant Professor of Psychiatry and Neurology, University of Pittsburgh, 3501 Forbes Ave, Pittsburgh, PA 15213 – USA. First author: Cécile Tissot, BSc, PhD candidate, McGill University, 6825 Boulevard LaSalle Montreal, QC H4H 1R3 – Canada, University of Pittsburgh, 3501 Forbes Avenue Pittsburgh, PA, 15213 – USA.

## Abstract

**Introduction:** [^18^F]MK6240 is a tau-PET tracer that quantifies brain tau neurofibrillary tangles (NFT) load in Alzheimer’s disease (AD). The aims of our study are to test the stability of common reference regions estimates in the cerebellum over time and across diagnoses and evaluate the effects of age-related and off-target retention in the longitudinal quantification of [^18^F]MK6240 in target regions.

**Methods:** We assessed reference, target, age-related and off-target regions in 125 individuals across the aging and AD spectrum with longitudinal [^18^F]MK6240 standardized uptake values (SUV) and ratios (SUVR) (2.25± 0.4 years of follow-up duration). We obtained SUVR values from meninges, a region exhibiting frequent off-target retention of [^18^F]MK6240, as well as compared tracer uptake between cognitively unimpaired young (CUY, mean age: 23.41± 3.3 years) and cognitively unimpaired older adults (CU, amyloid-β and tau negative, mean age: 58.50± 9.0 years) to identify possible, non-visually apparent, age-related signal. Two-tailed t-test and Pearson correlations tested the difference between groups and associations between changes in region uptake, respectively.

**Results:** Inferior cerebellar grey (CG) and full CG presented stable SUV cross-sectionally and over time, across diagnosis and Aβ status. [^18^F]MK6240 uptake was significantly different between CU young and adults mostly in putamen/pallidum (affecting ∼75% of the region) but also in Braak II region (affecting ∼35%), comprised of the entorhinal cortex and hippocampus. Changes in meningeal and putamen/pallidum SUVRs were not significantly different from zero, nor varied across diagnostic groups. We did not observe significant correlations between longitudinal changes in age-related or meningeal off-target retention and changes in target regions, whereas changes in all target regions were highly correlated.

**Conclusion:** Inferior and full CG were similar across diagnostic groups cross-sectionally and stable over time, and thus were deemed suitable reference regions for quantification. Despite this not being visually perceptible, [^18^F]MK6240 has age-related retention in subcortical regions, in much lower magnitude but topographically co-localized with the most significant off-target signal of the first-generation tau tracers. The lack of correlation between changes in age-related/meningeal and target retention suggests little influence of possible off-target signals on longitudinal tracer quantification. On the other hand, the age-related tracer retention in Braak II needs to be further investigated. Future post-mortem studies should elucidate the source of the newly reported age-related [^18^F]MK6240 signal, and *in vivo* studies should further explore its impact on tracer quantification.

## Introduction

Accumulation of amyloid-β (Aβ) plaques and hyperphosphorylated tau, forming neurofibrillary tangles (NFT), are hallmarks of Alzheimer’s disease (AD)(*1*) and can be observed in aging and AD dementia(*2*). Assessment of the tau levels in the brain is done through cerebrospinal fluid and positron emission tomography (PET) imaging. Radiotracers used in PET imaging are considered optimal when they present desirable characteristics such as rapidly equilibrating *in vivo* kinetics, low off-target retention, no significant lipophilic radiolabeled metabolites able to enter the brain and high affinity for their target(*3*).

[^18^F]MK6240 is a promising tracer allowing for the quantification of fibrillar tau pathology *in vivo* with post-mortem studies confirming its binding to paired helical fragments of phosphorylated tau(*4*–*7*). The tracer binds with high affinity to NFT, thus making it specific for AD-related tauopathy. As shown in post-mortem data, the tracer does not seem to bind to tau aggregates in non-AD tauopathies(*5,8*), except in rare frontotemporal dementia mutations associated with brain deposition of NFT(*9*). [^18^F]MK6240 allows for the differentiation between cognitively unimpaired (CU), mild cognitive impairment (MCI), and AD subjects(*4,10*). Furthermore, [^18^F]MK6240 has been shown to recapitulate *in vivo* the tau pathologic stages, proposed via post-mortem studies by Braak and colleagues(*11,12*).

Despite several favorable features of [^18^F]MK6240, some common challenges in PET studies remain unaddressed for this tracer, such as the choice of a reference region for longitudinal studies and the impact of off-target retention on tracer quantification in target regions (i.e. regions expected to show specific, tau-related retention of [^18^F]MK6240). Post-mortem and *in vivo* studies have indicated that [^18^F]MK6240 has off-target retention in neuromelanin-containing cells(*5*). Those are regions also observed using first-generation tau-PET tracers such as the substantia nigra (*13*). However, there is a much greater off-target retention of [^18^F]MK6240 in the meninges(*4,10*), and it is currently the main concern for an accurate quantification of NFT using this tracer.

As longitudinal tracer quantification is critical for clinical trials using tau-PET imaging agents as a possible surrogate marker of tau accumulation, exploring the optimal reference region and the effects of off-target retention on longitudinal [^18^F]MK6240 quantification is crucial(*14,15*). Here, we studied longitudinal changes in reference, target, age-related, and off-target regions across diagnostic groups and Aβ status to elucidate the caveats associated with the longitudinal quantification of [^18^F]MK6240.

## Methods

### Participants

We included individuals from the TRIAD cohort(*16*), with data obtained from December 2017 to November 2021. The study was approved by the Douglas Mental Institute Research Board and all participants gave written consent. Detailed information gathered from the participants can be found here: https://triad.tnl-mcgill.com/. All participants underwent a complete neuropsychological evaluation, a magnetic resonance imaging (MRI), and acquisition of both [^18^F]AZD4694 (Aβ) and [^18^F]MK6240 (tau) PET scans. We used two distinct subject samples for the analyses described in this work. To assess age-related off-target retention of [^18^F]MK6240, we included 36 cognitively unimpaired young (CUY - <35 years) and 28 cognitively unimpaired older adults (CU – 40-65 years), both presenting no AD-related pathology (Aβ and tau); this sample was called “age-related sample” and only included cross-sectional data. Aβ status was determined as [^18^F]AZD4694 global PET SUVR lower than 1.55 SUVR(*17*), while tau status was determined with [^18^F]MK6240 temporal meta-ROI lower than 1.24 SUVR, as previously described (*18*). The longitudinal sample was composed of 125 individuals [11 CUY, 66 CU Aβ negative (Aβ-), 17 CU Aβ positive (Aβ+), and 31 cognitively impaired (CI) Aβ+] who underwent a follow-up assessment between 1.5 and 3.5 years after their baseline. Baseline diagnosis was used in the analyses, following clinical assessments as well as Mini-Mental State Examination (MMSE), and Clinical Dementia Rating (CDR) scores and the NIA-AA criteria(*19*). CU individuals did not have an objective impairment, MMSE score of 26 or more and CDR score of 0(*20*). Individuals diagnosed with mild cognitive impairment (MCI) had subjective and/or objective cognitive impairment, relatively preserved activities of daily life, as assessed with an MMSE score of 26 or above and CDR of 0.5(*21*). Dementia due to AD was assessed with an MMSE score of less than 26 and a CDR score of 0.5 or more. No participant met the criteria for another neurological or major neuropsychiatric disorder following a clinical interview performed by a trained physician.

### PET image processing

Participants underwent a T1-weighted MRI (3T, Siemens), as well as [^18^F]MK6240 tau-PET and [^18^F]AZD4694 Aβ-PET using the same brain-dedicated Siemens high resolution research tomograph. [^18^F]MK6240 images were acquired 90-110 minutes post tracer injection and reconstructed using an ordered subset expectation maximization (OSEM) algorithm on a 4D volume with four frames (4 × 300 s)(*4*). [^18^F]AZD4694 images were acquired 40-70 minutes post tracer injection, and reconstructed with the same OSEM algorithm with three frames (3 × 600s)(*4*). Each PET acquisition was finished with a 6-min transmission scan with a rotation ^137^Cs point source for attenuation correction. Images were further corrected for motion, decay, dead time, and random and scattered coincidences. Standardized uptake value (SUV) images were calculated considering the injected radionuclide dose and weight of each participant 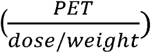. SUV values were extracted from the inferior cerebellar grey (CG), superior CG, Crus I, and full CG. SUV ratio (SUVR) images were generated using the inferior CG as the reference region for [^18^F]MK6240 and full CG for [^18^F]AZD4694. Finally, images were spatially smoothed to achieve an 8-mm full-width at half-maximum resolution. Meninges were not masked at any step of the processing for the present study. A populational-based meningeal mask was created with the Montreal Neurological Institute (MNI) MINC-toolkit as the region having >90% of probability of being part of either telencephalon or cerebellar meninges in CUY individuals (Supplementary Figure 1). SUVR values in Braak regions were extracted following Pascoal *et al*(*11*). Additionally, the Desikan-Killiany-Tourville (DKT) atlas(*22*) was used to obtain [^18^F]MK6240 SUVR from the putamen and the pallidum. A global [^18^F]AZD4694 SUVR value was estimated by averaging the SUVR from the precuneus, prefrontal, orbitofrontal, parietal, temporal, anterior, and posterior cingulate cortices(*23*). The cut-off value for Aβ positivity used is a published threshold of 1.55(*17*) global SUVR, applied to classify participants as Aβ positive (Aβ+) or Aβ negative (Aβ-). MNI MINC-toolkit was used to calculate average images of [^18^F]MK6240 retention.

### Statistical analyses

R statistical software (version 4.0.0) was used to perform non-imaging statistical analyses. T-tests or ANOVA tests were conducted for continuous variables and chi-square or Fisher tests for categorical variables for demographic information when appropriate. Longitudinal change (Δ) was calculated as follows: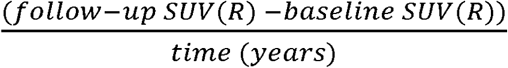. Associations between changes in biomarkers were assessed with Pearson correlation. Voxel-wise statistical comparisons were conducted using VoxelStats, a statistical toolbox implemented in MATLAB(*24*). The age-related retention was evaluated at the voxel level conducting a two-sided t-test, between CUY and CU Aβ and tau negative elderly individuals aged from 35 to 65 years (Table 1.A). False discovery rate correction was applied with a voxel-level correction of *p* < 0.05.

**Table 1:**
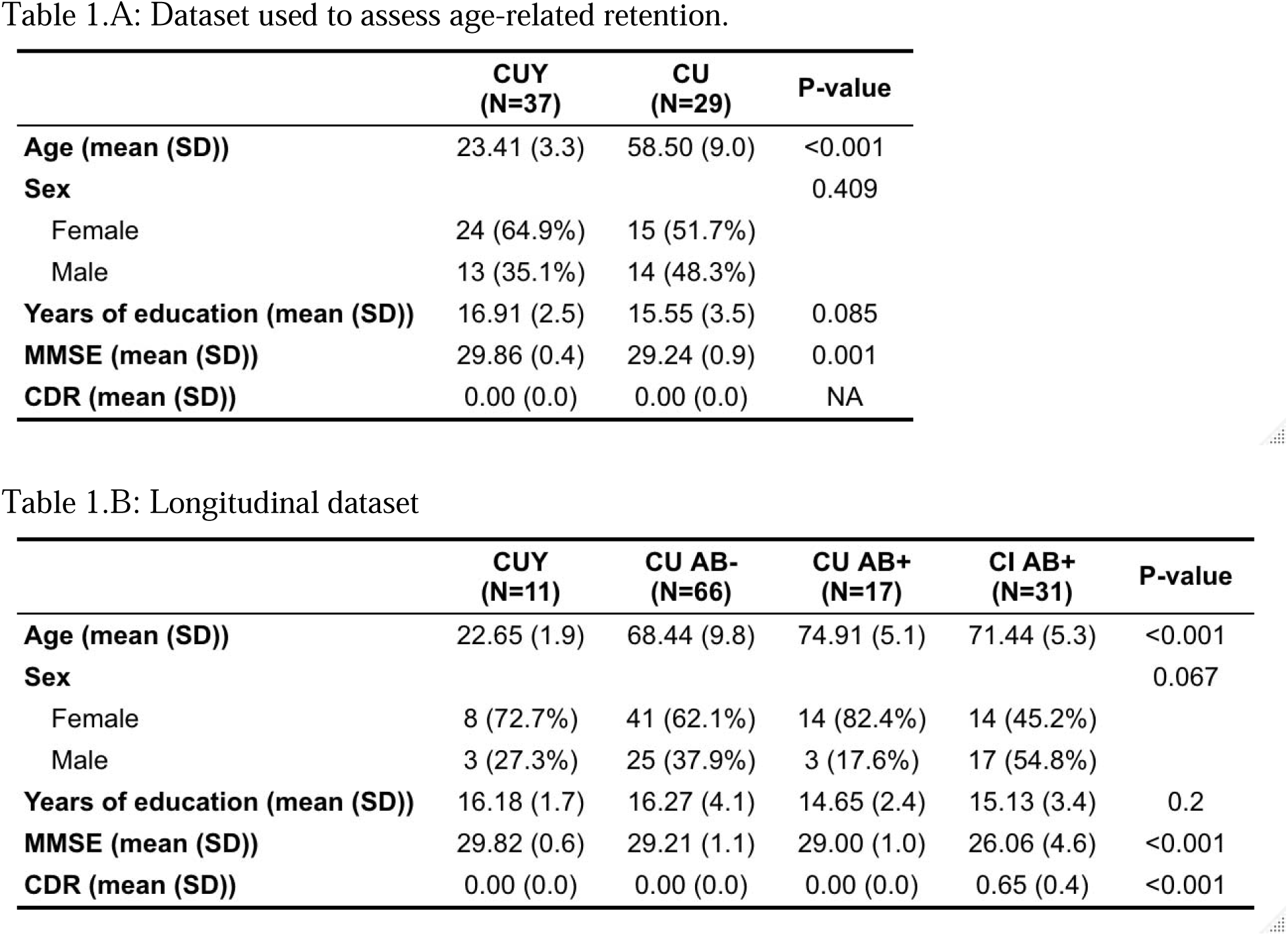
Demographics

|  | **CUY**<br>**(N=37)** | **CU**<br>**(N=29)** | **P-value** |
| --- | --- | --- | --- |
| **Age (mean (SD))** | 23.41 (3.3) | 58.50 (9.0) | <0.001 |
| **Sex** |  |  | 0.409 |
| Female | 24 (64.9%) | 15 (51.7%) |  |
| Male | 13 (35.1%) | 14 (48.3%) |  |
| **Years of education (mean (SD))** | 16.91 (2.5) | 15.55 (3.5) | 0.085 |
| **MMSE (mean (SD))** | 29.86 (0.4) | 29.24 (0.9) | 0.001 |
| **CDR (mean (SD))** | 0.00 (0.0) | 0.00 (0.0) | NA |

|  | **CUY**<br>**(N=11)** | **CU AB-**<br>**(N=66)** | **CU AB+**<br>**(N=17)** | **CI AB+**<br>**(N=31)** | **P-value** |
| --- | --- | --- | --- | --- | --- |
| **Age (mean (SD))** | 22.65 (1.9) | 68.44 (9.8) | 74.91 (5.1) | 71.44 (5.3) | <0.001 |
| **Sex** |  |  |  |  | 0.067 |
| Female | 8 (72.7%) | 41 (62.1%) | 14 (82.4%) | 14 (45.2%) |  |
| Male | 3 (27.3%) | 25 (37.9%) | 3 (17.6%) | 17 (54.8%) |  |
| **Years of education (mean (SD))** | 16.18 (1.7) | 16.27 (4.1) | 14.65 (2.4) | 15.13 (3.4) | 0.2 |
| **MMSE (mean (SD))** | 29.82 (0.6) | 29.21 (1.1) | 29.00 (1.0) | 26.06 (4.6) | <0.001 |
| **CDR (mean (SD))** | 0.00 (0.0) | 0.00 (0.0) | 0.00 (0.0) | 0.65 (0.4) | <0.001 |

## Results

### Participants

In the age-related group analyses, comparing CUY and CU older adults who were both Aβ and tau negative, we observed no significant difference in sex and years of education. By definition, subjects had a significant difference in age. We also observed a small but significant difference in the MMSE score, with the CU older adults having a slightly lower score (Table 1.A). In the longitudinal dataset, as expected, we observed significant differences in the age, MMSE and CDR scores across groups. There was no difference in the years of education; however, a small but significant difference was observed regarding sex with more females in the CUY and CU Aβ+ groups (Table 1.B).

### Assessment of stability of reference regions over time for use in longitudinal studies

Our first objective was to ascertain the reference region appropriateness for longitudinal quantification of [^18^F]MK6240. Using the change of SUV over time (ΔSUV), we tested the stability of SUV over the time frame of our study in the inferior CG, superior CG, cerebellar Crus I, and full CG. No significant differences in the longitudinal changes were observed when separating individuals based on their clinical diagnosis (Figure 1.A), Aβ status (Figure 1.B), or both (Figure 1.C). Coefficients of variation of [^18^F]MK6240 ΔSUV were similar with the highest numerical value in the inferior CG longitudinal change (−12.64), whereas the Crus I presented the lowest (−3.705) (Figure 1.D, Table 2.A). No significant variability in SUV value was found in any one of the assessed reference regions (Table 2.B).

**Figure 1:**
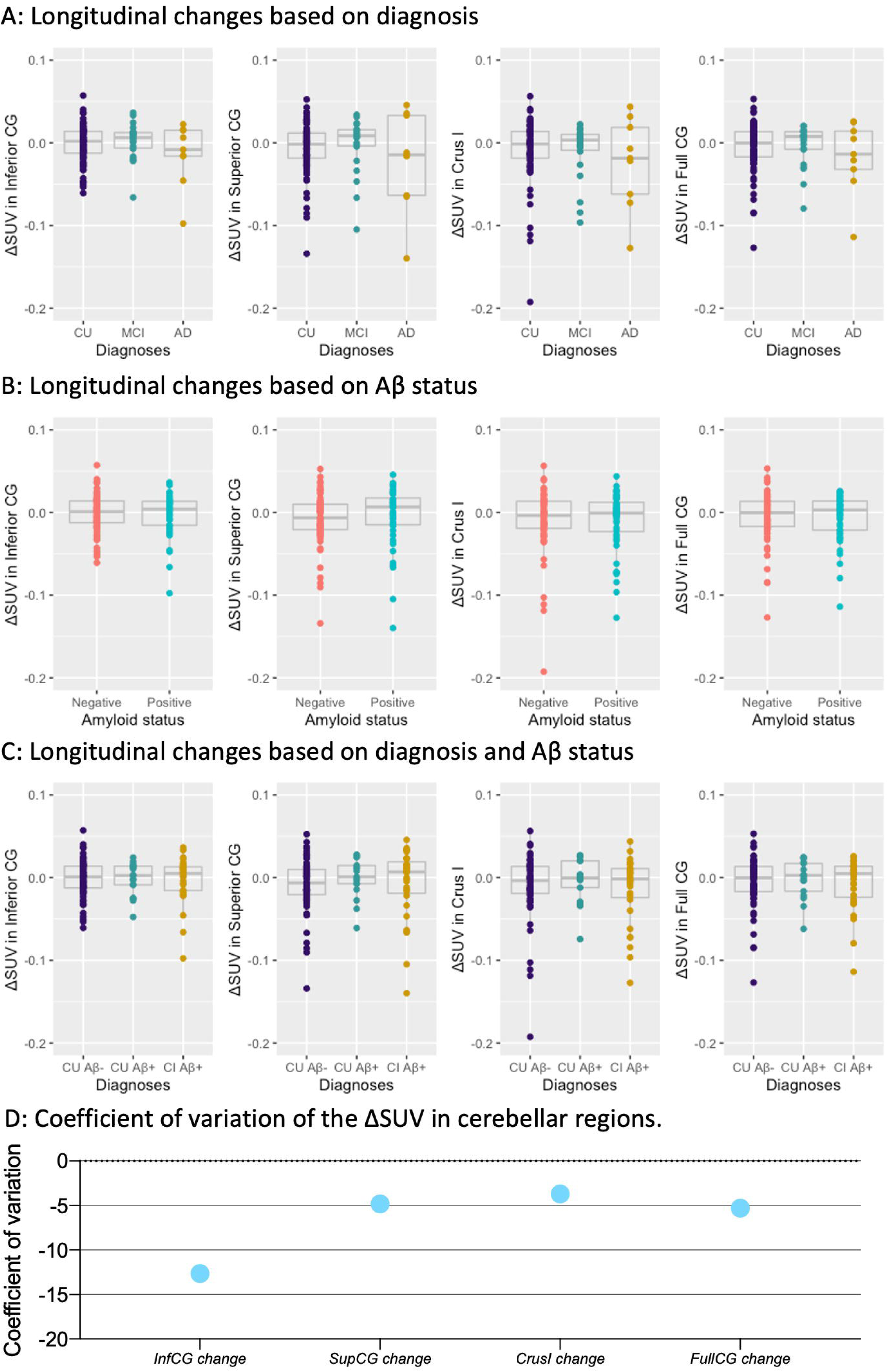
Annualized longitudinal changes in [^18^F]MK6240 SUV in cerebellar candidate reference regions. ΔSUV of [^18^F]MK6240 was not significantly different across A) diagnosis, B) Aβ status, C) diagnosis and Aβ status. D) Coefficient of variation of longitudinal changes in SUV within reference regions. Δ = change calculated as 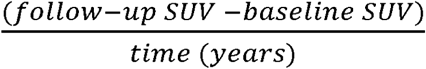.

**Table 2:** Coefficient of variation and differences in cerebellar regions SUV based on diagnosis

|  | **All** | **CU Aβ-** | **CU Aβ+** | **CI Aβ+** |
| --- | --- | --- | --- | --- |
| *InfCG BL* | 0.185 | 0.181 | 0.141 | 0.217 |
| *SupCG BL* | 0.272 | 0.238 | 0.186 | 0.323 |
| *CrusI BL* | 0.253 | 0.256 | 0.193 | 0.284 |
| *FullCG BL* | 0.219 | 0.221 | 0.186 | 0.237 |
| *InfCG FU* | 0.172 | 0.179 | 0.161 | 0.165 |
| *SupCG FU* | 0.248 | 0.208 | 0.209 | 0.288 |
| *CrusI FU* | 0.224 | 0.212 | 0.219 | 0.252 |
| *FullCG FU* | 0.197 | 0.201 | 0.207 | 0.188 |
| *InfCG change* | -12.644 | -14.347 | -11.122 | -11.031 |
| *SupCG change* | -4.828 | -3.791 | -52.033 | -5.05 |
| *CrusI change* | -3.705 | -3.836 | -7.484 | -2.930 |
| *FullCG change* | -5.297 | -5.468 | -9.630 | -4.281 |

|  | **All** | **CU Aβ-** | **CU Aβ+** | **CI Aβ+** |
| --- | --- | --- | --- | --- |
| *InfCG* | 0.674 | 0.812 | 0.720 | 0.830 |
| *SupCG* | 0.269 | 0.211 | 0.943 | 0.662 |
| *CrusI* | 0.086 | 0.178 | 0.722 | 0.310 |
| *FullCG* | 0.246 | 0.400 | 0.756 | 0.457 |

Supplementary material displays the cross-sectional differences in those SUV values. In the cross-sectional analysis, only superior CG (CU-AD *p*-value <0.001, MCI-AD *p*-value <0.001) and Crus I (CU-AD *p*-value = 0.046, MCI-AD *p*-value = 0.051) presented significant differences between diagnostic groups after correction for multiple comparisons (Supplementary Figure 2). Cross-sectional coefficients of variation of reference regions are reported in Supplementary Figure 3.

### Meningeal and age-related retentions

Figure 2.A represents the average [^18^F]MK6240 SUVR values in the CUY group. We observed no significant difference between diagnostic groups in the telencephalon meninges cross-sectionally [CU Aβ- vs CU Aβ+ *p*-value = 0.891; CU Aβ- vs CI Aβ+ *p*-value = 0.797; CU Aβ+ vs CI Aβ+ *p*-value = 0.999] and longitudinally [CU Aβ- vs CU Aβ+ *p*-value = 0.150; CU Aβ- vs CI Aβ+ *p*-value = 0.677; CU Aβ+ vs CI Aβ+ *p*-value = 0.524]. Similarly, no differences were observed in the cerebellar meninges neither cross-sectionally [CU Aβ- vs CU Aβ+ *p*-value = 0.946; CU Aβ- vs CI Aβ+ *p*-value = 0.919; CU Aβ+ vs CI Aβ+ *p*-value = 0.837] nor in longitudinal changes [CU Aβ- vs CU Aβ+ *p*-value = 0.563; CU Aβ- vs CI Aβ+ *p*-value = 0.631; CU Aβ+ vs CI Aβ+ *p*-value = 0.963] (Figure 2.B).

**Figure 2:**
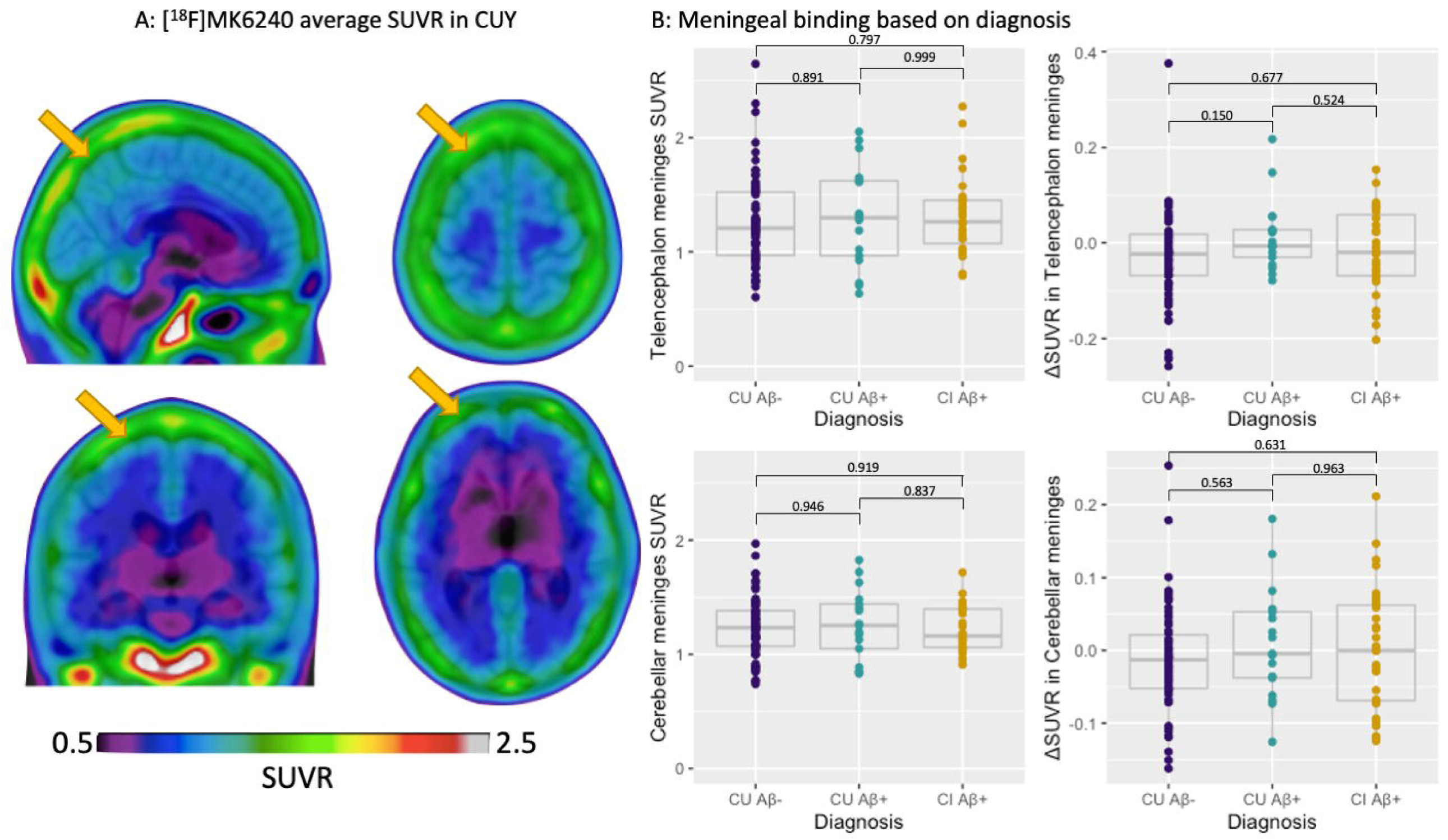
Cross-sectional and longitudinal meningeal [^18^F]MK6240 SUVR across groups. A) Representative [^18^F]MK6240 average SUVR image in cognitively unimpaired young individuals. B) Cross-sectional and longitudinal changes in [^18^F]MK6240 (ΔSUVR) in the telencephalon and cerebellar meninges did not indicate significant differences depending on diagnosis and Aβ status. Δ = change calculated as 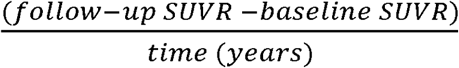.

Average [^18^F]MK6240 SUVR images of CUY and CU Aβ and tau negative individuals did not seem to display readily apparent visual differences. However, a t-test between the two groups revealed significantly higher [^18^F]MK6240 retention in the putamen, the pallidum, a parcel of cerebellar white matter, as well as a few other cortical regions (Figure 3.A). We assessed the percentage of overlap between the age-related signal and brain regions. The most important regional overlaps were observed with the putamen (75% of the region showing overlap), pallidum (72%), followed by Braak stage II region (38%) (Figure 3.B). SUVR values in the putamen and pallidum were significantly different in CI Aβ+ individuals as compared to CU (Aβ- and Aβ+) cross-sectionally [CU Aβ- vs CU Aβ+ *p*-value = 0.926; CU Aβ- vs CI Aβ+ *p*-value < 0.001; CU Aβ+ vs CI Aβ+ *p*-value < 0.001], but not in the longitudinal rate of change (ΔSUVR) [CU Aβ- vs CU Aβ+ *p*-value = 0.880; CU Aβ- vs CI Aβ+ *p*-value = 0.728; CU Aβ+ vs CI Aβ+ *p*-value = 0.994] (Figure 3.C).

**Figure 3:**
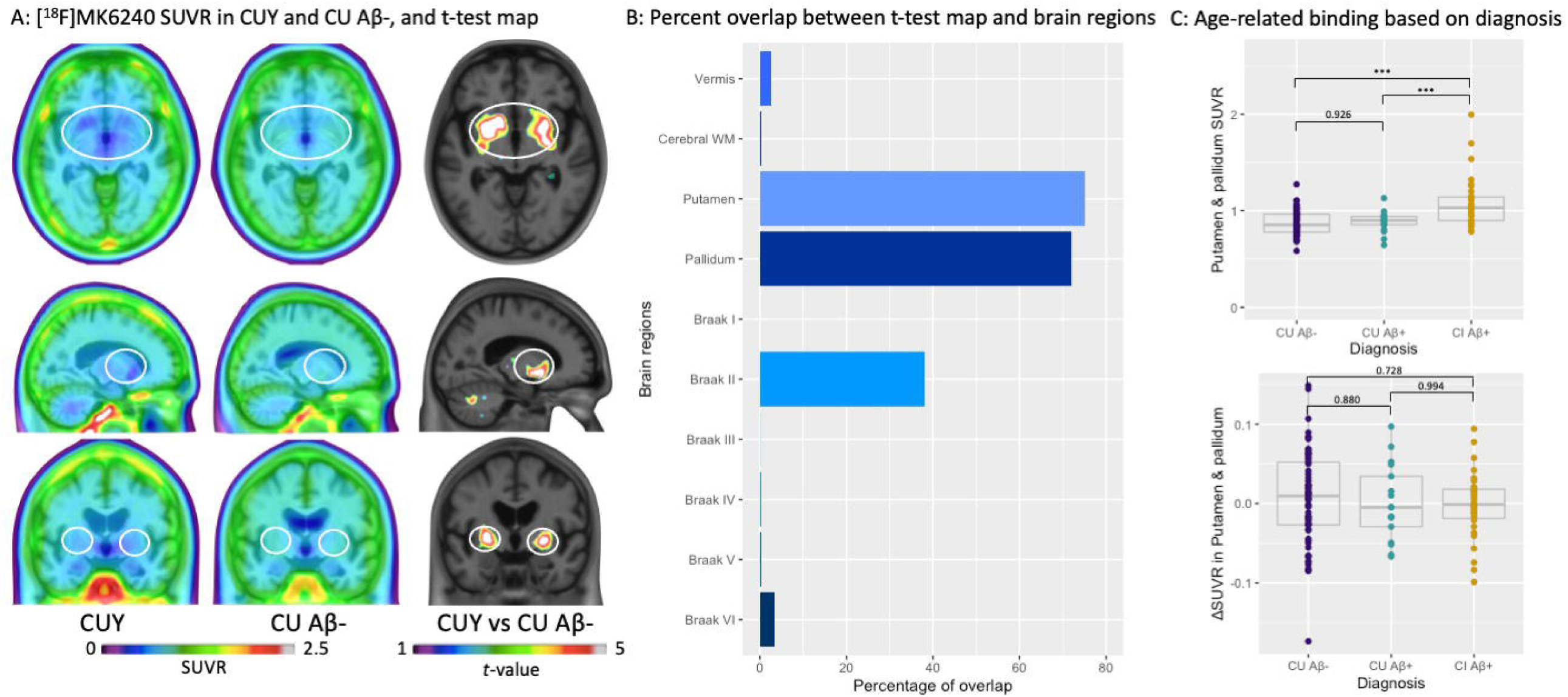
[^18^F]MK6240 age-related retention. A) [^18^F]MK6240 average SUVR image in cognitively unimpaired young (CUY) and cognitively unimpaired older adults (CU) Aβ negative individuals do not seem to show strong differences visually. T-test between the two groups depicts age-related retention in the putamen, the pallidum, cortical regions, and the cerebellar white matter. B) Percentage of overlap between age-related t-map and anatomical brain regions. C) Longitudinal changes (ΔSUVR) in [^18^F]MK6240 SUVR values in the putamen/pallidum did not present significant differences across groups, whereas cross-section SUVR was higher in CI individuals. *P*-values portrayed as *p*<0.001=***. Δ = change calculated as 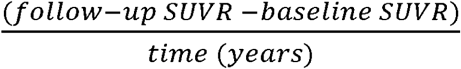.

### Associations of changes in target, age-related, and off-target retention

Target regions were considered brain regions in which we expect to see [^18^F]MK6240 retention based on the pattern of tau distribution extensively reported in the post-mortem literature(*1,25*). We found a strong correlation between ΔSUVR in target regions with each other, where each Braak region was more strongly correlated with the adjacent stages. The weakest correlation was depicted between Braak I and Braak VI regions. When extracting the average SUVR in Braak I-III and Braak IV-VI, we also observed a strong positive correlation between regions (R = 0.62, *p*-value < 0.001). We then used SUVR values in the telencephalon and cerebellar meninges, as well as the putamen and pallidum in the correlations. ΔSUVR in those regions did not correlate significantly with ΔSUVR in any one of the target regions [Braak I-III and telencephalon meninges: R = -0.031, *p*-value = 0.74; Braak IV-VI and telencephalon meninges: R = 0.1, *p*-value = 0.29; Braak I-III and cerebellar meninges: R = -0.12, *p*-value = 0.2; Braak IV-VI and cerebellar meninges: R = -0.015, *p*-value = 0.87; Braak I-III and putamen & pallidum: R = 0.12, *p*-value = 0.2; Braak IV-VI and putamen & pallidum: R = 0.05, *p*-value = 0.6]. Nor did the meningeal and putamen/pallidum ΔSUVR correlate with each other [R = 0.0099, *p*-value = 0.92] (Figure 4).

**Figure 4:**
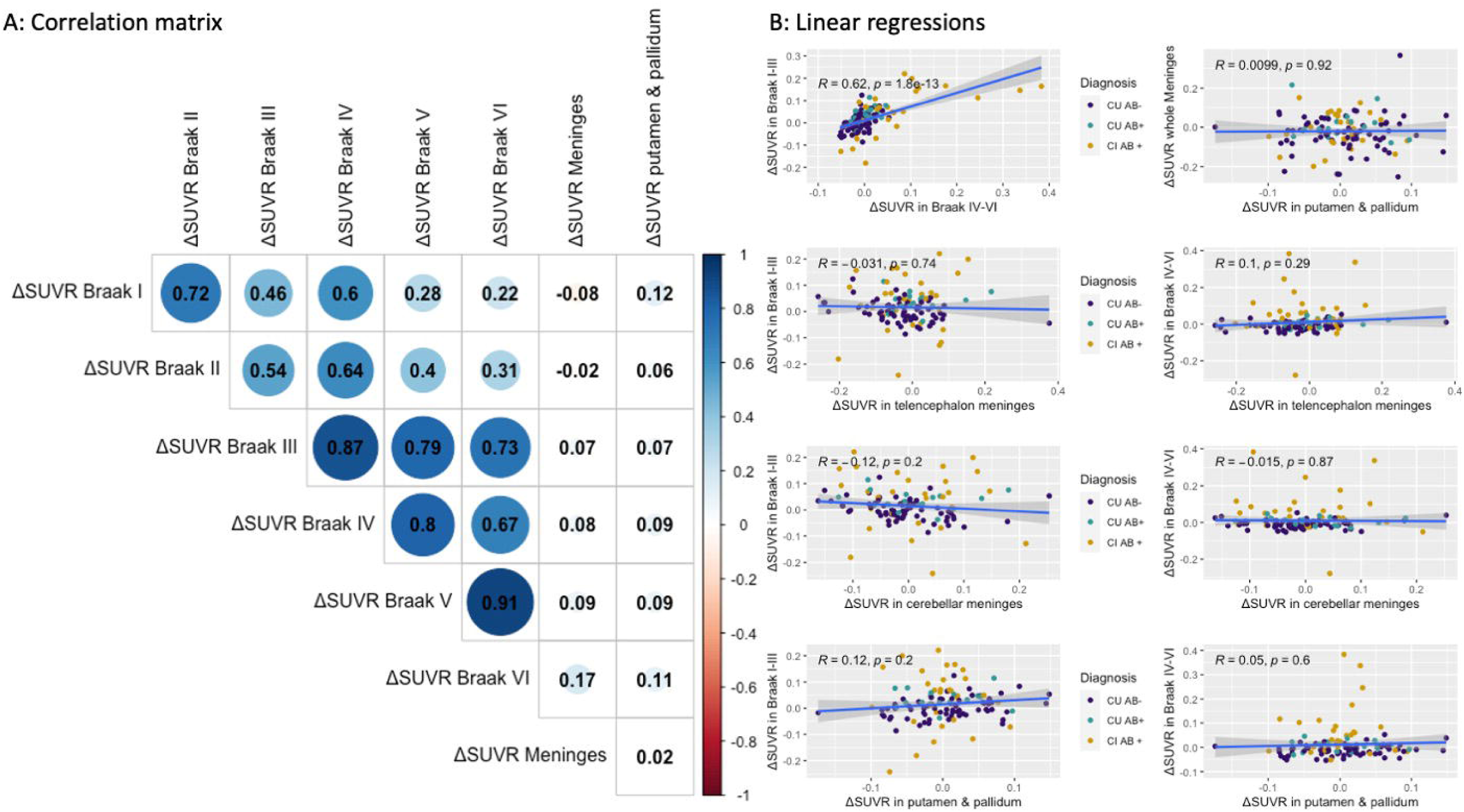
Correlations between longitudinal changes in SUVR in age-related, meningeal, and target regions. We assessed ΔSUVR values in regions presenting target (Braak I-VI), off-target (both telencephalon and cerebellar meninges), and age-related (putamen/pallidum) tracer uptake. We observed strong correlations between ΔSUVR among target regions; however, those did not correlate with ΔSUVRs in off-target and age-related regions. A) The matrix and B) plots present the estimates of Pearson correlations between regions. Δ = change calculated as 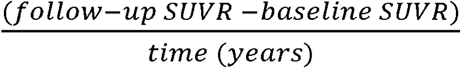.

## Discussion

This study suggests that the most widely used cerebellar reference regions (inferior CG, superior CG, Crus I, and full CG) present stability, with no changes over time, and therefore may be suitable for use in longitudinal studies, although differences were observed in superior CG and Crus I cross-sectionally. We provide evidence for the existence of an age-related retention in the putamen/pallidum, similar, albeit of much lower magnitude, to the reported off-target retention observed using the first-generation tau-PET tracers(*13,26*). Finally, we demonstrated that there was no association between [^18^F]MK6240 ΔSUVR in target regions and in age-related or meningeal off-target signal over the time frame of our study.

Previous cross-sectional studies have already shown that indices of tau load made using [^18^F]MK6240 are amenable to simplified tissue ratio methods using data acquired 90-110 minutes post-injection(*4,7*). However, questions remain regarding the suitability of the reference region for longitudinal tracer quantification due to the bias often inherent to SUVR data(*10,11*). This is of paramount importance because [^18^F]MK6240 has been used in clinical trial settings to capture longitudinal changes in tau tangle pathology(*27*). Extensive research has been conducted focusing on the cerebellum as the appropriate reference region for tau radiotracers(*28,29*), as the CG is not expected to harbor significant NFT pathology (*1*). Although the gold standard method for the assessment of the optimal reference region relies on dynamic quantifications with arterial input function, one crucial characteristic of a reference region for longitudinal assessments is not having large variability over time, across diagnostic groups or other pathological features (i.e., Aβ status in the case of AD research)(*30*). In this study, we estimated the [^18^F]MK6240 SUV values in distinct regions of the cerebellar grey matter at baseline, as well as its change over time, based on diagnosis and on Aβ status. Results indicate that there were small cross-sectional differences between diagnostic groups in the superior CG and Crus I but not in the inferior CG and full CG. Additionally, we did not observe differences in the cerebellar SUVs based on Aβ status. When examining the differences between baseline and follow-up assessments, we did not observe any significant difference in [^18^F]MK6240 SUV for any cerebellar region. Even though all variability was relatively minor, we observed the lowest numerical variability in the SUV levels of the inferior CG cross-sectionally and in the Crus I longitudinally. Altogether, our results suggest that tracer retention in the tested reference regions was relatively stable over time and across diagnostic groups, suggesting that all these reference regions could potentially be used for longitudinal [^18^F]MK6240 quantification. Given the cross-sectional differences in superior CG and Crus I but not in the inferior CG and full CG, these latter regions seem the most appropriate ones for the computation of [^18^F]MK6240 SUVR values. Future studies using the gold standard arterial input function should address other important characteristics of an optimal reference region.

The t-test comparing young individuals under age 35 and participants between 40 and 65 years of age allowed us to assess age-related retention of [^18^F]MK6240. The regions presenting the higher age-related [^18^F]MK6240 retention were the putamen and pallidum. Those are often considered off-target regions using other radiotracers for tau(*29*–*31*), but it seems to be of lower magnitude with [^18^F]MK6240(*32*). As we included participants younger than 65 years that were CU Aβ and tau negative, we do not expect on-target [^18^F]MK6240 retention in subcortical structures based on the post-mortem literature(*33*). Indeed, subcortical regions have been shown to harbor NFT accumulation only at late Braak pathological stages (*25,34*), which might explain why we observed a significant cross-sectional difference in putamen and pallidum SUVR between CI Aβ+ individuals as compared to CU (either Aβ- or Aβ+). While the tracer retention observed in Braak II can represent an age-related signal, we cannot entirely exclude that there is some true concentration of NFT in this region, as modest tau accumulation in CU Aβ-individuals has already been reported in the hippocampus (*2,33*). Another possibility is that the marked off-target retention of first-generation tracers in the choroid plexus, which contaminates the Braak II region for these tracers, could be a minor age-related problem with [^18^F]MK6240 as well. Finally, we assessed meningeal retention in both telencephalon and cerebellar regions, which has already been characterized as off-target by previous post-mortem studies(*5*). Importantly, we observed no significant difference in the magnitude of meningeal uptake across diagnosis and Aβ status and no change over time. Taken together, those results suggest that besides the meningeal retention, [^18^F]MK6240 presents a newly described age-related retention in subcortical brain regions, in which the cause(s) need to be elucidated by future *in vitro* studies across the aging spectrum.

We observed no association between annualized [^18^F]MK6240 SUVR changes in target, age-related regions, and meninges. Braak regions were used to represent target areas for tau tangles, as extensively reported in post-mortem studies(*1,25*). ΔSUVR values in target brain regions correlated strongly with each other, suggesting that changes in tracer retention in these brain regions are influenced by the same brain process, likely NFT accumulation(*36*). On the other hand, we did not observe a significant correlation between ΔSUVR in meningeal off-target uptake and changes in Braak target regions. Additionally, age-related ΔSUVR in the putamen and pallidum did not correlate with that in target regions. Nor did off-target, meningeal and age-related subcortical ΔSUVR correlate with each other. This suggests that different processes set the paces of progression in target, off-target, and age-related regions and that the spillover from off-target regions would not heavily influence rates of progression in [^18^F]MK6240 target regions.

This study has limitations. The lack of arterial sampling at baseline and follow-up limits the assessments of reference region and accurate tracer retention. Post-mortem data would validate our findings, as it would allow us to ensure the absence of NFT in the cerebellar regions, as well as in the age-related retention regions. Without post-mortem confirmation, we cannot exclude that age-related retention in CU older adults is caused by tau tangle pathology. Moreover, our sample was restricted to a follow-up time of 1.5 to 3.5 years; by using other follow-up durations, we might obtain different results. In the current project, we evaluated only a small subset of reference regions that are frequently reported in the literature; it is possible that other regions might present better results for [^18^F]MK6240 longitudinal quantification. An additional limitation is that we did not provide any evidence about the mechanism through which age-related retention occurs. Arterial and post-mortem data are needed to understand our findings.

## Conclusion

Inferior CG and full CG are suitable reference regions for the cross-sectional and longitudinal quantification of [^18^F]MK6240. [^18^F]MK6240 exhibits off-target retention in the meninges and an age-related signal in the putamen/pallidum, also likely representing off-target retention, and Braak II region in which the source needs to be elucidated. The lack of an association between changes in SUVR within age-related, off-target, and target regions suggests that longitudinal changes in [^18^F]MK6240 are not heavily driven by changes in age-related or off-target signals. However, future post-mortem studies are needed to clarify these findings.

## Data Availability

All data produced in the present study are available upon reasonable request to the authors.

