## Supplementary Material for "The association of age-related and off-target retention with longitudinal quantification of [^18^F]MK6240 tau-PET in target regions"

Supplementary figures:

**Supplementary Figure 1**: Probabilistic mask used to calculate telencephalon (red) and cerebellar (green) meningeal retention.

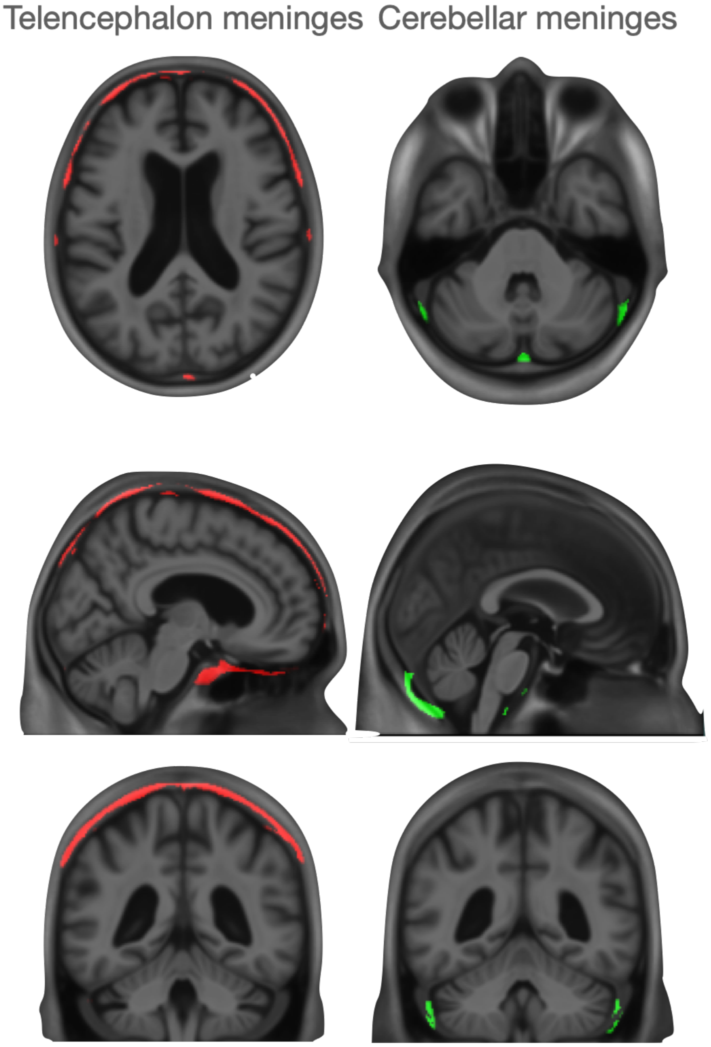


This mask was defined as region having >90% probability to present high retention in CU individuals.

**Supplementary Figure 2:** [$^{18}$F]MK6240 SUV values across different cerebellar regions measured cross-sectionally.

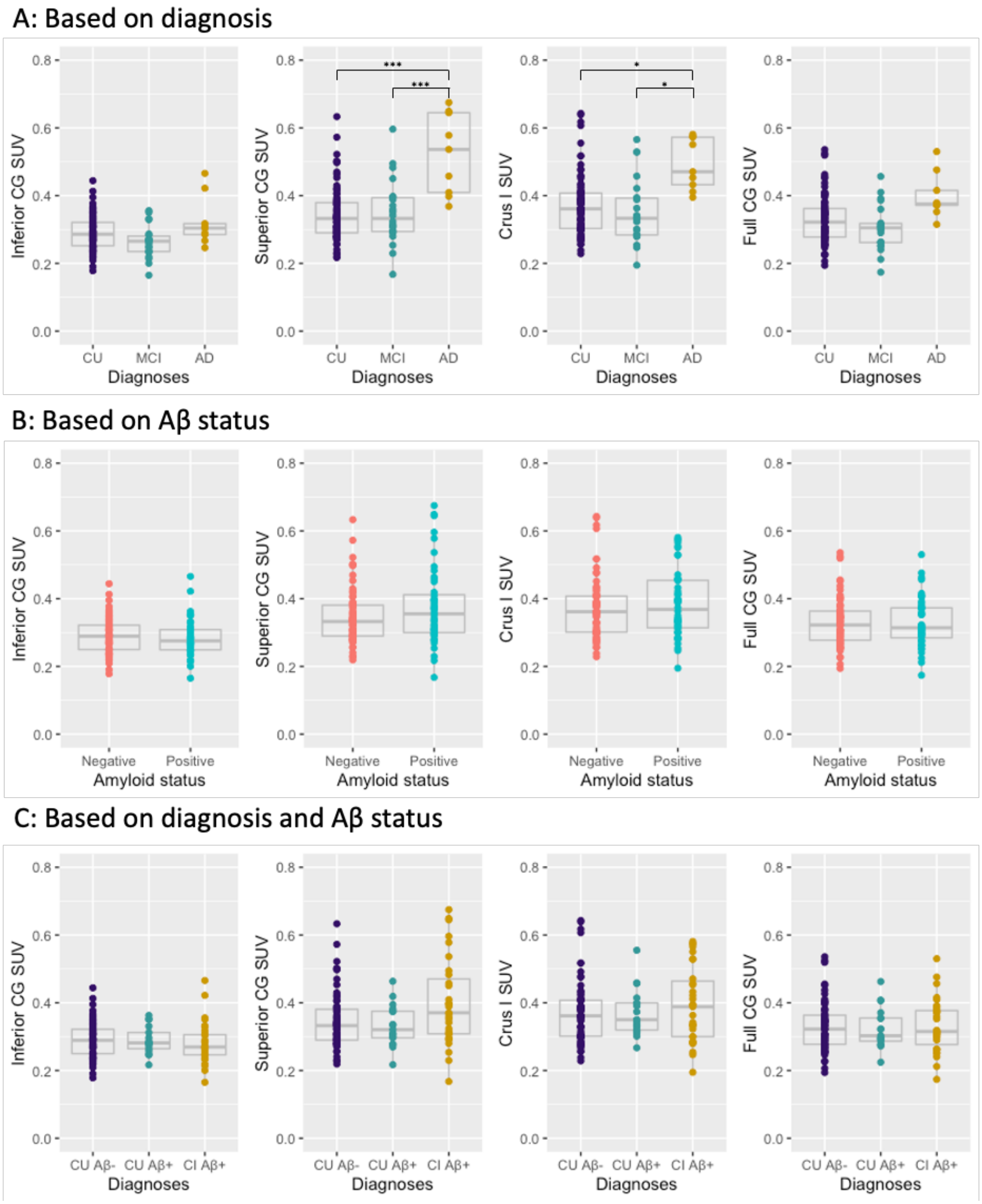

Cross-sectional [[18]F]MK6240 SUV values did not demonstrate significant differences based on A) diagnosis, B) Aβ status C) diagnosis and Aβ status. *P*-values portrayed as *p*-value < 0.001=***, *p*-value < 0.5=*.

**Supplementary Figure 3:** Cross-sectional coefficient of variation of [[18]F]MK6240 SUV in cerebellar regions, measured at baseline or at follow-up visit.

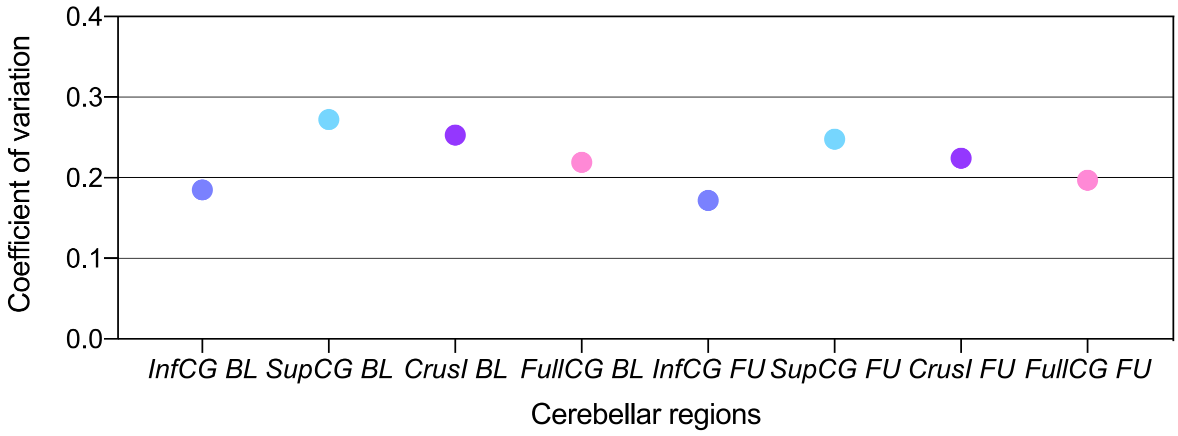
